# Elevated pre-treatment Neutrophil-to-lymphocyte and neutrophil-to-lymphocyte-platelet ratios are associated with a high risk of in-hospital death among patients with prostate cancer in Nigeria

**DOI:** 10.1101/2023.08.11.23293979

**Authors:** Jude Ogechukwu Okoye, Vivian Ifunanya Ogbonnaya, Michael Emeka Chiemeka, Samuel Ifedioranma Ogenyi

## Abstract

**Introduction:** In the last decade, there is an increasing mortality rate among patients diagnosed with prostate cancer (PCa) in West Africa. To identify the causes of the high mortality rate, this study analyzed the occurrence of high-grade tumours and the presence of BRCA2 mutation. It also assessed the systemic inflammatory indices as prognostic tools in low-resource settings.

**Methods:** This study included 72 cases of PCa diagnosed from Jan. 2017 to Dec. 2020. The neutrophil-to-lymphocyte ratio (NLR), platelet-to-lymphocyte ratio (PLR), platelets-neutrophils-to-lymphocytes ratio (PNLR), and neutrophils-to-lymphocytes platelets ratio (NLPR) were assessed and analyzed accordingly. Significance was set at p< 0.05.

**Results:** The prevalence of Gleason grades (G) 1 to 5 was 9.3%, 16.3%, 16.3%, 25.6, and 32.6%, respectively. There was a high frequency of BRCA2 mutation (58.3%) and the frequency was higher among patients with G4/5 tumours (59.5%) than in patients with G1-G3 tumours (46.7%) at p= 0.347. A high frequency of G4/5 tumours was observed among patients within the age group of 50-59 years (n= 7/8; 87.5%) and patients with castration-resistant PCa (n= 12/17; 70.6%). The pre-treatment PLR and calcium concentration were higher among patients with G4/5 tumours compared to patients with G1-G3 tumours (p= 0.046 and < 0.001, respectively.) There were direct relationships between BRCA2 expression and age (p= 0.019), tumour grade and calcium (p= 0.000), BRCA2 and calcium expression (p= 0.027), unemployment and G4/5 (p< 0.001), and education status and G4/5 (p= 0.020). The pretreatment NLR and NLPR were 2.0 and 4.7 times higher in in-hospital deaths than in stable discharges at p= 0.005 and 0.001, respectively.

**Conclusion:** This study revealed high frequencies of BRCA2 mutation and high-grade PCa in Southern Nigeria. It also suggests that pre-treatment PLR could be used to identify patients with G4/5 while pretreatment NLR and NLPR could be used to predict treatment outcomes.

## Introduction

Globally, prostate cancer (PCa) is the third most common [1]. The odds of developing PCa are 1 in 52 and 1 in 9 for countries with low and high sociodemographic index (SDI), respectively [2]. However, the age-standardized incidence rate (ASIR) to age-standardized mortality rate (ASMR) is 2.4 times higher in countries with low SDI than in countries with high SDI [1]. Felay et al. also reported that PCa is the most common cancer and the leading cause of cancer-related deaths among males in West Africa [3]. From 2018 and 2020, the ASIR and ASMR increased by 1.2% and 1.6% in West Africa, respectively [1,4]. The risk for developing PCa includes age, smoking, high BMI, family history of malignancy, BRCA1/2 mutations, and Lynch syndrome [1]. Although studies show that rapid disease progression and higher mortality are higher among BRCA2 mutation carriers compared with BRCA1 mutation carriers [5,6], other reasons for the increase or variation in incidence and mortality rates are yet to be elucidated and extensively investigated. Identifying individuals at risk of death using a less invasive and affordable approach could improve the survival rates of patients diagnosed with PCa in West Africa. Inflammatory response involving neutrophils, lymphocytes and platelets plays a crucial role in tumour initiation and metastasis [7]. In Asia, high pre-treatment systemic immune inflammation indices (SIII) are associated with ethnicity, tumour type, poor overall survival, and poor progression-free survival [8]. Man and Chen also reported that high neutrophil-to-lymphocyte ratio (NLR) and platelet-to-lymphocyte ratio (PLR) are associated with poor prognosis, especially among patients with metastatic castration resistance PCa [9]. In this study, we assessed SIII in high and low-grade prostate tumours as well as in-hospital deaths and patients stable on discharge to identify prognostic tools among Nigerian men. For the first time, this study showed that NLPR could be used as a prognostic tool, especially among patients with low-grade tumours.

## Methods

### Study Population and Ethics

This retrospective study included 72 cases of prostate cancer diagnosed from January 2017 to December 2020 at the Department of Gynaecology, Nnamdi Azikiwe University Teaching Hospital (NAUTH), Nnewi, Nigeria. Only two patients received docetaxel as platinum chemotherapy. This study was approved by the NAUTH ethics committee (NAUTH/CS/66/VOL.14/VER.3/116/2021/078). The patient’s medical records were accessed for socio-clinical demographics such as age, gender, comorbidities, and presentation time. All analyses were performed by the ethical standards in the Declaration of Helsinki.

### Sample collection and handling

Two samples, 5 ml of veinous whole blood, were collected from each patient and discharged into EDTA containers: a week before the first chemotherapy and a week before discharge. Full blood counts were carried out on the whole blood samples using a Haemo-autoanalyzer. Following ultrasound investigations, biopsy, and surgery, resected tissues were sent to the Department of Morbid Anatomy and Forensic Medicine for histological investigation. Two pathologists evaluated the tissues for evidence of malignancy based on the Gleason pattern of grading (G). The total white cell count (TWBC) (10^^9^/L), neutrophil-to-lymphocyte ratio (NLR), platelet-to-lymphocyte ratio (PLR), platelets-neutrophils to lymphocytes ratio (PNLR; [Platelet count x Neutrophil count]/Lymphocyte count), and neutrophil-to-lymphocyte-platelet ratio (NLPR; [Neutrophil count x 100]/Lymphocyte count x platelet count]) were calculated for the subgroups.

### Procedure for Immunohistochemistry

The sections were first dewaxed and hydrated. The Epitopes in sections were then retrieved. Sections were treated with peroxidase blocker and subsequently washed in phosphate Buffered Saline (PBS). The sections were treated with the primary antibody (Breast Cancer gene 2; BRCA2) for 60 minutes in a humidity chamber, washed in PBS, and treated with the secondary antibody accordingly. The slides were then washed in PBS for 2 minutes. The sections were treated with Horseradish peroxidase and washed in 2 changes of PBS. The sections were stained with 3,3′-Diaminobenzidine (1 drop in 1 ml of Substrate), washed in PBS, stained with Haematoxylin, washed in PBS, and distilled water, dehydrated, cleared, and mounted with a DPX. BRCA2 staining intensity or expression in the tissues was scored using a scale of 0, +1, +2, and +3. For BRCA2 and Iron expressions, scores 0 and +1 were considered negative while scores +2 and +3 were considered positive.

### Statistical analysis

Chi-square/Fisher was used to determine the association between the socio-clinical demographics of patients 50 years and those > 50. Pearson’s correlation was used to determine the relationship between the SIII before and after the last treatment. T-test was used for comparing data of 1. Patients aged ≤ 50 years and > 50 years, 2. Chemotherapy naïve and experienced patients, 3. Patients who received 1-3 cycles and 4-6 cycles of chemotherapy, 4. Herbal medicine experience and naïve patients, and 5. Patients with and without metastatic tumours. ANOVA was used to compare data of patients who presented at ≤ 6 months and > 6 months, and patients who were stable, unstable, and dead at discharge (in-hospital death). Significance (p) was set at p≤ 0.050.

## Result

### Socio-clinical features of PCa cohort

In this study, the frequency of PCa peaked in the age group of 70-79 years (figure 1). The prevalence of Gleason grades 1 to 5 was 9.3%, 16.3%, 16.3%, 25.6, and 32.6%, respectively. The mean (± SD) age of patients with Gleason grades 1 to 5 was 70.3 ± 6.9, 77.3 ± 6.2, 75.6 ± 12.6, 72.8 ± 7.8, and 67.4 ± 9.8, respectively (p= 0.191). There was an inverse relationship between tumour grade and age (p= 0.108). A higher frequency of G4/5 tumours (n= 7/8; 87.5%) was observed among patients aged 50-59 years (p> 0.05). Most of the patients were self-employed, and a higher frequency of G4/5 (high-grade) tumours was observed among patients who were unemployed (p< 0.05). Most of the patients had only basic education and the frequency of high-grade PCa decreased with increasing education (p< 0.05). The frequency of G4/5 tumours was also high among patients with a history of alcohol and tobacco use, and positive for DM (p> 0.05). Patients who were positive for BRCA2 mutation and herbal-experienced had a higher and a lower frequency of high-grade tumours, respectively (p> 0.05 and < 0.05).

**Figure 1:**
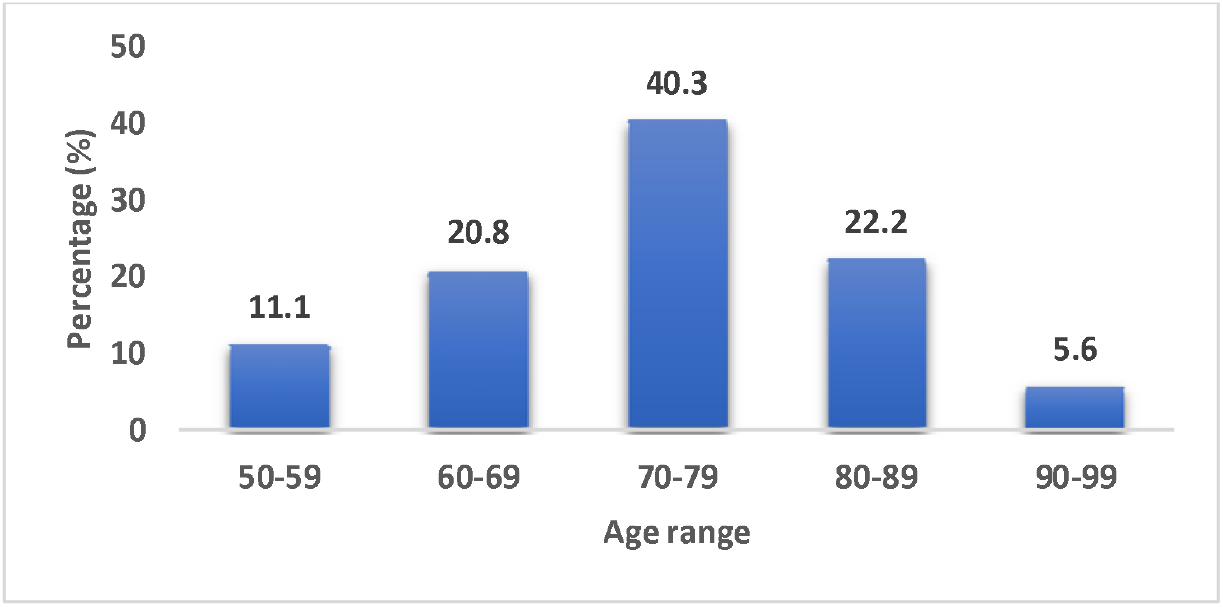
Frequency of prostate cancer across age groups Figure 1 shows a high frequency of early-onset PCa among patients aged 50-59 years.

The frequency of high-grade tumours was higher among patients who presented within 6 months of symptom manifestation (p> 0.05). Among men who had bilateral orchidectomy, the frequency of castration-resistant PCa (CRPCa) was 47.2% (17/36). A higher frequency of grade 4/5 tumours was observed among patients with CRPCa (70.6%; 12/17) compared with their castration-sensitive PCa (42.1%; 8/19) counterparts (p= 0.315). Approximately 6% of the patients received chemotherapy (table 1). The prevalence of BRCA2-negative and positive tissues was 65.1% and 34.9%, respectively (figure 2) whereas the prevalence of calcium-negative and positive tissues was 48.7% and 51.3%, respectively. The frequency of high-grade tumours was higher than G1-G3 (low-grade) and the age of the former was lower than the latter at p< 0.05 and > 0.05, respectively. The expression of BRCA2 and calcium was higher in high-grade than in low-grade at p> 0.05 and < 0.05, respectively. The lymphocyte and platelet counts were 1.3 times and 1.2 times higher while the neutrophil count was 1.2 lower in high-grade than in low-grade tumours at p= 0.122, 0.342, 0.054, respectively. There were direct relationships between BRCA2 expression and age (p= 0.019), Calcium and age (p= 0.281), tumour grade and Calcium (p= 0.000), and BRCA2 and Calcium expressions (p= 0.027). Higher frequencies of BRCA2 and calcium positivity were observed in high-grade compared with low-grade at p< 0.001.

**Table 1:** Socio-clinical characteristics of patients diagnosed with PCa.

| Variables | Total<br>N= 72 | G4/5<br>n= 42 (%) | p- value |
| --- | --- | --- | --- |
| **Level of Education:** |  |  |  |
| No formal education | 10 (13.9) | 7 (70.0) | 0.020* |
| Basic education | 57 (79.2) | 35 (61.4) |  |
| Tertiary education | 5 (6.9) | 0 (0.0) |  |
| **Employment status:** |  |  |  |
| Civil servant | 19 (26.4) | 3 (15.8) | < 0.001* |
| Self employed | 40 (55.5) | 29 (72.5) |  |
| Unemployed | 13 (18.1) | 10 (76.9) |  |
| **Alcohol consumption:** |  |  |  |
| No | 25 (34.7) | 12 (48.0) | 0.329 |
| Yes | 47 (65.3) | 30 (63.8) |  |
| **Tobacco Use:** |  |  |  |
| No | 40 (55.6) | 23 (57.5) | > 0.999 |
| Yes | 32 (44.4) | 19 (59.4) |  |
| **History of Hypertension:** |  |  |  |
| No | 33 (45.8) | 20 (60.6) | 0.812 |
| Yes | 39 (54.2) | 22 (56.4) |  |
| **Diabetes Mellitus** |  |  |  |
| No | 54 (75) | 30 (55.6) | 0.582 |
| Yes | 18 (25) | 12 (66.7) |  |
| **History of Herbal medicine:** |  |  |  |
| No | 45 (62.5) | 31 (68.9) | 0.027* |
| Yes | 27 (37.5) | 11 (40.7) |  |
| **TSMP** |  |  |  |
| ≤ 6 months | 41 (56.9) | 25 (61.0) | 0.636 |
| > 6 months | 31 (43.1) | 17 (54.8) |  |
| **BRCA2 mutation** |  |  |  |
| Negative | 30 (41.7) | 17 (56.7) | 0.815 |
| Positive | 42 (58.3) | 25 (59.5) |  |
| **Chemotherapy experience:** |  |  |  |
| No | 68 (94.4) | 38 (55.9) | 0.135 |
| Yes | 4 (5.6) | 4 (100) |  |
| **Bilateral Orchiectomy** |  |  |  |
| No | 36 (50.0) | 19 (50.0) | 0.474 |
| Yes | 36 (50.0) | 23 (67.6) |  |

Keys: TSMP= Time of symptom manifestation to presentation. Descriptive analysis and Chi-square/Fisher's exact test. \*Significance was set at $p \leq 0.05$ .

**Figure 2:**
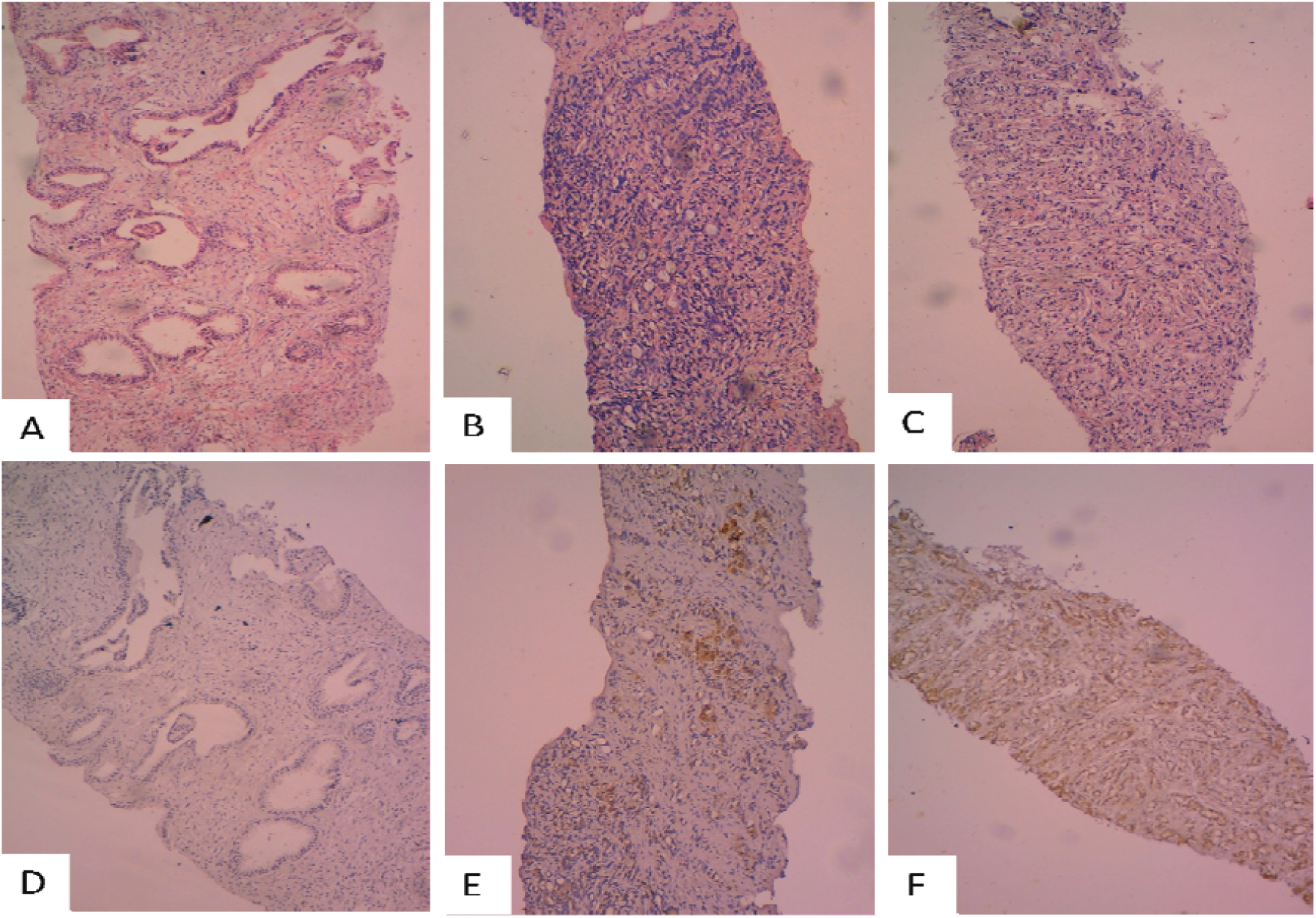
Sections of Prostate cancer tissues. Figures A-C were stained by the H&E technique while figures D-F were stained by the immunohistochemical technique. Figures D (control), E and F show negative, mild, and high staining intensity for BRCA2 protein, respectively. X200 magnification.

### Variation of Haematological indices based on socio-clinical characteristics

The median value of NLR, PLR, PNLR, NLPR, TPSA, and %FPSA of the patients was 2.27, 123.5, 434.8, 1.29, 129.7, and 10.20, respectively while their mean ± SD was 4.22 ± 1.13, 137.2 ± 16.27, 911.5 ± 306.10, 2.36 ± 0.59, 391.10 ± 26.00, and 12.09 ± 2.26, respectively. Pre-treatment PLR was higher among patients with high-grade tumours compared to low-grade at p< 0.05 whereas lower pre-treatment TWBC, NLR, PNLR, and NLPR were lower among the former than the latter at p< 0.05, p> 0.05, > 0.05 and < 0.05, respectively (figure 3). The pre-treatment TWBC, PLR, NLR, PNLR, and NLPR were 1.1, 1.1, 1.2, 1.2, and 1.7 times higher among patients who presented at the clinic after 6 months of symptom manifestation compared to patients who presented within 6 months of symptom manifestation (p> 0.05). The pre-treatment TWBC/PLR of herbal medicine-experienced patients was 1.2 and 1.7 times higher than that of herbal medicine-naïve patients (7.62 ± 1.27/194.2 ± 38.69 vs 6.13 ± 0.38/116.0 ± 10.83 at p= 0.161 and 0.019, respectively). The NLPR is 1.7 times higher among patients with CRPCa compared to patients with castration-sensitive PCa (0.94 ± 0.17 vs 1.59 ± 0.39, respectively; p= 0.087). The pre-treatment NLR and NLPR were 2.0 and 4.7 times higher in in-hospital deaths (IHD) than in stable discharges at p< 0.05 (table 2). Interestingly, PLR, TWBC, and PNLR were 0.8, 0.9, and 0.9 times lower in in-hospital deaths than in stable discharges (p> 0.05).

**Table 2:**
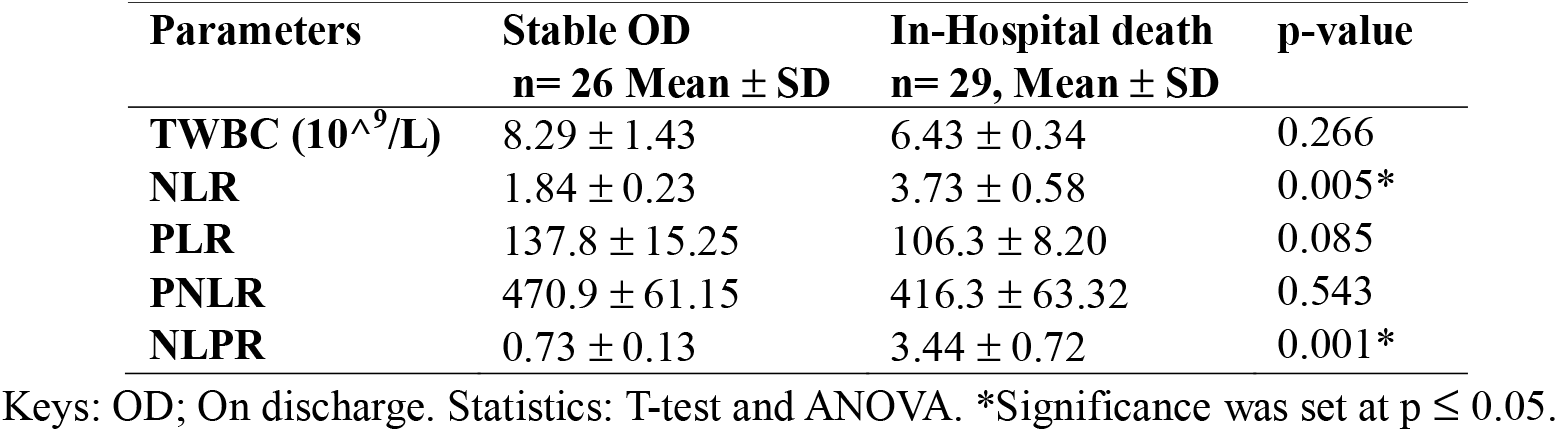
Comparative analysis of Haematological indices based on patient’s condition on discharge.

| Parameters | Stable OD<br>n= 26 Mean ± SD | In-Hospital death<br>n= 29, Mean ± SD | p-value |
| --- | --- | --- | --- |
| TWBC ($10^9/L$) | 8.29 ± 1.43 | 6.43 ± 0.34 | 0.266 |
| NLR | 1.84 ± 0.23 | 3.73 ± 0.58 | 0.005* |
| PLR | 137.8 ± 15.25 | 106.3 ± 8.20 | 0.085 |
| PNLR | 470.9 ± 61.15 | 416.3 ± 63.32 | 0.543 |
| NLPR | 0.73 ± 0.13 | 3.44 ± 0.72 | 0.001* |

Keys: OD; On discharge. Statistics: T-test and ANOVA. \*Significance was set at $p \leq 0.05$ .

**Figure 3:**
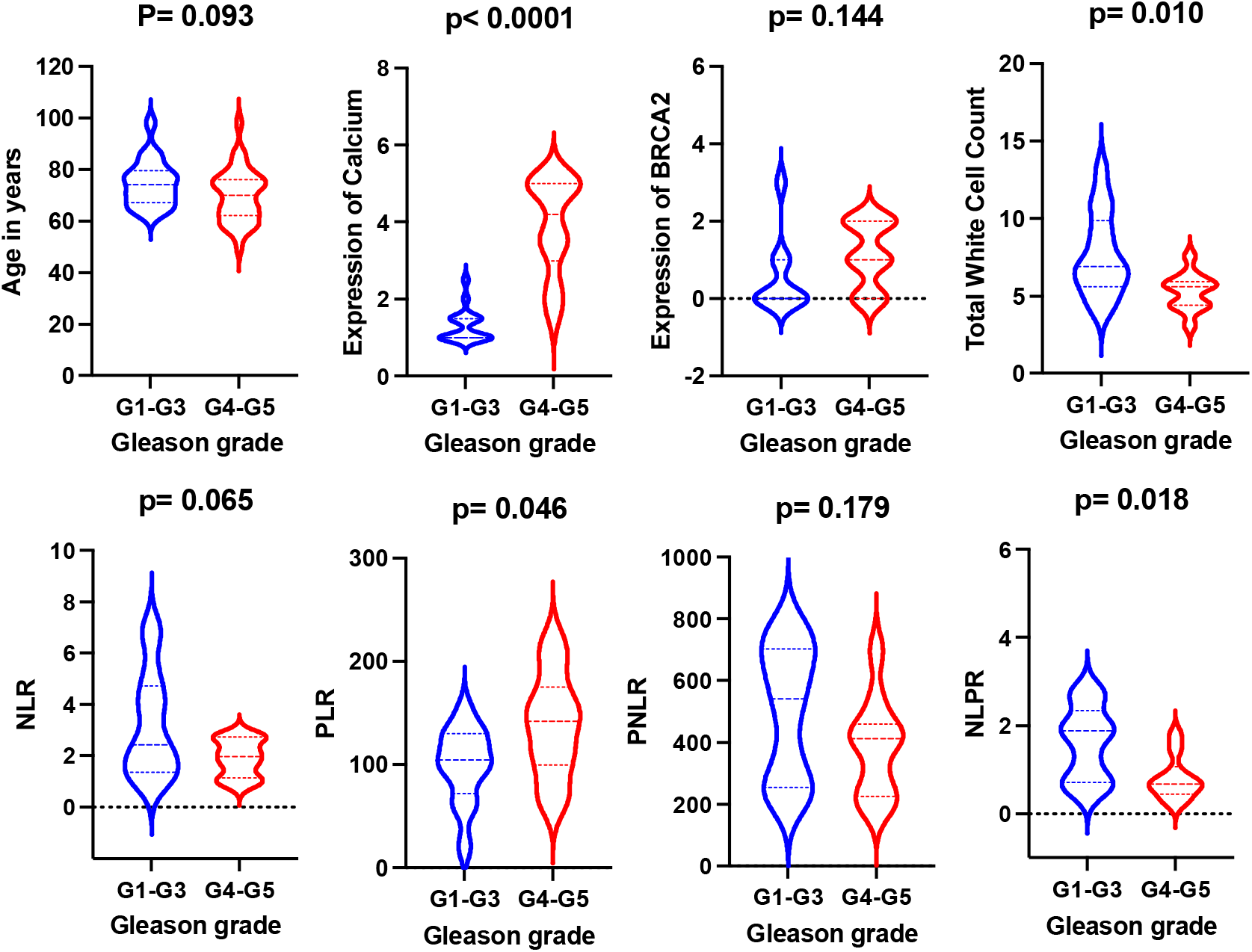
Comparison of age, Calcium and SIII profile based on tumour grade. Figure 3 shows a higher frequency of early-onset G4-G5 tumours marked by high PLR, and high expression of calcium and BRCA2 protein at p< 0.05, < 0.05 and > 0.05. It also revealed a high TWBC, NLR, NLPR and PNLR among patients with G1-G3 (low-grade) tumours than in patients with G4-G5 (high-grade) tumours at p< 0.05, < 0.05, < 0.05 and > 0.05, respectively.

## Discussion

To identify affordable biomarkers for identifying individuals at risk of PCa-related death, this study evaluated the pre-treatment NLR, PLR, PNLR and NLPR in different clinical outcomes. The frequency of PCa in the age group of 50-59 years in this study is lower than the pooled frequency of 18.4% recorded among West African men [2]. The difference may be attributed to improved awareness and uptake of screening exercises. The frequency of high-grade tumours in this study is also higher than the pooled frequency (48.8%) earlier recorded in West Africa [2]. It is also higher than the frequencies recorded in the United States of America (USA; 13.0%) and Asia (25.3%) [10]. Evidence shows that germline mutation in the BRCA2 gene is a major predictive factor of aggressive prostate cancer and poor survival [6]. This might explain why there was a higher frequency of high-grade tumours among patients with BRCA2 mutation in this study. The frequency of BRCA2 gene mutation in this study is also higher than the frequency recorded among African Americans (14.3%) and Caucasians (3.6%) [10]. Taken together, the high ASMR in West Africa could be attributed to high BRCA2 mutation and high-grade tumours. It could be argued that the BRCA2 mutation was responsible for the defective calcium (Ca^2+^) channel resulting in the high concentration of calcium in high-grade tumours [11]. The defective Ca^2+^ channel has been linked to malignant transformation, tumour proliferation, metastasis, and treatment resistance [12]. This also suggests that calcium concentration in prostate tissues could be used for solving cases of intra-observer variability.

For the first time, this study showed that NLPR could be used as a prognostic tool, especially among patients with low-grade tumours. In this study also, a high NLR was observed in IHD cases even though only 33.3% of IHD were associated with grade high-grade tumours. There was also an insignificant difference in NLR between low-grade and grade high-grade tumours. This is supported by an earlier study that did not show any significant association between the Gleason score and the pathological stage [13]. This suggests that NLR is more strongly associated with treatment outcomes than diagnosis. This is supported by two meta-analyses which show that elevated NLR predicts poor prognosis and could be used for risk stratification among patients with PCa [14,15]. The lower PLR observed in IHD compared to stable discharge, and the high PLR among patients with high-grade tumours compared to low-grade tumours suggests PLR could be used to monitor disease progression. This is at variance with the findings of Huszno et al. who did not observe any significant association between PLR and clinicopathological factors among men with PCa in Poland [16].

## Conclusion

This study revealed a high prevalence of high-grade and aggressive PCa in Nigeria. This might be an explanation for the high mortality rate among West African men who were diagnosed with the disease. The study also revealed elevated pretreatment NLR and NLPR among patients with poor outcomes and suggests that they offer predictive advantages in low-resource settings where cancer care is borne by the patients. The study suggests that patients aged 50-59 years should be closely monitored for improved survival.

## Data Availability

All data produced in the present study are available upon reasonable request to the authors

